# Community engagement for COVID-19 prevention and control: A Rapid Evidence Synthesis

**DOI:** 10.1101/2020.06.17.20133876

**Authors:** Brynne Gilmore, Rawlance Ndejjo, Adalbert Tchetchia, Vergil de Claro, Elizabeth Nyamupachitu-Mago, Alpha A Diallo, Claudia Abreu Lopes, Sanghita Bhattacharyya

## Abstract

**Introduction:** Community engagement has been considered a fundamental component of past outbreaks, such as Ebola. The COVID-19 pandemic and its control efforts require social actions and behaviours, all of which place a large reliance on individual and community compliance, highlighting the need for appropriate community engagement to support such work. However, there is concern over the lack of involvement of communities within COVID-19 thus far. Identifying how community engagement approaches have been used in past epidemics may support more robust implementation of community engagement within COVID-19 response.

**Methodology:** A rapid evidence review was conducted to identity how community engagement is used for infectious disease prevention and control during epidemics. Three databases (PubMed, CINHAL and Scopus) were searched in addition to extensive snowballing for grey literature. Previous epidemics were limited to Ebola, Zika, SARS, MERS and H1N1 since 2000. No restrictions were applied to study design or language, though articles must have detailed a minimum of one community engagement for infection prevention and control initiative. All authors participated in searching, screening, and data extraction, with a minimum of two authors at each stage.

**Results:** From 1,112 references identified in our search, 32 articles met our inclusion criteria. All but 3 articles were published on or after 2015 which details 37 community engagement initiatives for Ebola (n=28), Zika (n=5) and H1N1 (n=4). Twenty-seven of these initiatives were implemented in low-income countries and 10 from high-income countries. Six broad community engagement actors were identified: local leaders, community and faith-based organisations, community groups, health facility or community health committees, individuals and key stakeholders. These actors worked across six different functions: designing and planning, community entry and trust-building, social and behaviour change communication, risk communication, surveillance and tracing, and logistics and administration. Leaders were the most prevalent actor being engaged, and behaviour change communication, risk communication, and surveillance and tracing were the most common function of community engagement. Implementation considerations community engagement in prevention and control of COVID-19 are reported within.

**Conclusion:** COVID-19’s global presence and social transmission pathways require social and community responses. This may be particularly important to reach marginalised populations and support equity-informed responses. Previous experience from outbreaks shows that community engagement can take many forms and include different actors and approaches who support various prevention and control activities. Countries worldwide are encouraged to assess existing community engagement structures, and utilise community engagement approaches to support contextually specific, acceptable and appropriate COVID-19 prevention and control measures.

## Introduction

Community engagement within health is crucial to achieve primary health care and promote people-centred services [1-3]. It can support buy-in and sustainability of health interventions [4], health advocacy [5], improved quality and satisfaction of services [6], and contribute to health systems responsiveness [7] and strengthening [8]. Community engagement has more recently been considered a fundamental component during outbreaks, largely arising during the 2014-2015 Ebola epidemic in West Africa.

The way people interact and live with each other through their structures as well as their historical pathways require considerations on how to effectively adapt and respond to any disease outbreak. For example, differences in political-cultural and social structures, systems and processes amongst communities affect health behaviours and outcomes during disease outbreaks [9]. Experience with public health emergencies of international concern highlight the need for contextually appropriate community engagement strategies [10-16]. Moreover, a recent rapid review noted key lessons in risk communication for control of outbreaks to include communities taking a central role in the response, involving local leaders and groups, tailoring interventions to communities and ensuring a two-way communication [12].

Early implementation of prevention and control activities during the 2014-2015 Ebola epidemic had several barriers, including suspicions regarding the existence of the disease and motives of the government and international organisations [10, 14]. To address these barriers community engagement became a key pillar to the response. Several measures to engage communities were undertaken, including building partnerships with local and religious leaders and working with the community to develop and adjust key messages on behavioural changes [10, 17]. These measures significantly contributed to the success achieved in controlling the outbreak and ensuring the resilience of the health system [10, 17, 18].

In relation to COVID-19, community engagement can be critical for creating local and context specific solutions to prevention and control responses [19]. Through this “bottom-up approach”, communities participate in “decision-making processes of planning, design, governance and delivery of services aimed at improving population health and reducing health inequalities” [18]. The COVID-19 pandemic as a total social phenomenon should include actively engaging and adapting local views, voices and concerns in health crisis response efforts [19]. Moreover, the World Health Organization’s recommended measures to prevent and control COVID-19 such as physical-social distancing, case identification and contact tracing require understanding of the different social dynamics in communities and how these can better be leveraged to minimize the impact of the epidemic [20, 21]. The measures have a huge reliance on communities reigniting the importance of community engagement to build trust and delay disease spread as drug and vaccine development efforts continue.

However, there is concern over the lack of involvement of communities within COVID-19. Rajan and colleagues note limited number of WHO member states reporting to have a COVID-19 community engagement plans [22]. The scientific community -mainly drawn by social scientists-has called for the attention of funders and implementers on the relevance of community engagement for COVID-19 [23-26], with other international stakeholders, including WHO, UNICEF and IFRC echoing its importance [27]. Recent reviews on global evidence for COVID-19 have focused on community health workers [28] providing important evidence and insights to guide response. However, there is no evidence synthesis that addresses how community engagement can be utilised for COVID-19 prevention and control. Thus, we conducted a rapid evidence review on community engagement for infectious disease prevention and control to learn lessons for COVID-19 and future pandemic response.

## Review focus

This review wanted to understand ‘how community engagement is used for infectious disease prevention and control during epidemics. In doing so, we specifically aimed to identify: what approaches and community actors are involved; what interventions are conducted; who are the target groups of community engagement and how are equity considerations incorporated; what are the linkages and relationship to other health system stakeholders; and what are main implementation considerations for successful community engagement for infectious disease prevention and control?

## Methodology

Given the emergency nature of the recent COVID-19 global pandemic, we conducted a rapid evidence review to support timely findings. Rapid reviews are a form of evidence synthesis that tailor the methodology of a systematic review to produce contextually relevant evidence on an arising topic in a timely and efficient manner [29]. To support the expedited nature of rapid reviews, several recommendations are put forth, including narrowing the scope, limiting the number of searches or electronic databases, using one reviewer for study screening and selection, and parallelisation of review tasks [29]. This rapid review followed the methodology suggested by the Alliance for Health Policy and Systems Research [30]. A co-production team comprising all authors of this paper was established through the collaborative platform ‘Collectivity’.

A protocol was developed and agreed upon by the research team, which comprises academics, implementers and policy makers from multiple disciplines and backgrounds, all members of the Community Health – Community of Practice of Collectivity. The team then conducted a rapid evidence review of academic and grey literature in May, 2020. The main focus of the review was to identify what types of community engagement approaches are used within infectious disease prevention and control, which required articles to describe a minimum of one specific initiative. As such, no criteria for effectiveness or outcomes were applied. Full inclusion and exclusion criteria can be found in Table 1.

**Table 1:** Inclusion and Exclusion Criteria

| Topic | Inclusion Criteria | Exclusion Criteria |
| --- | --- | --- |
| Intervention/<br>Population | Describes a specific Community engagement approach or activity | Community health worker programmes<br>Structures without community members serving the same community |
| Focus | Prevention and/or control of infectious diseases: Ebola, SARS, MERS, Zika and H1N1 | Not focused on prevention and/or control of infectious disease |
| Scope of intervention | Community level | Not community focused |
| Time | Published on or after 2000 | Published before 2000 |
| Article type | Primary, empirical studies, of any design, programme reports and descriptions that provide learning on specific CE approaches | Commentaries, abstracts. No specific CE approach detailed |
| Language | All languages included. Searching done in English and some French terms | No exclusion criteria |

For this review, we excluded Community Health Worker approaches and interventions as reviews of this nature have already been conducted [28] and we narrowed our scope to include five recent infectious disease outbreaks: Ebola, SARS, MERS, Zika and H1N1.

### Databases and Snowballing

In line with rapid review recommendations, we limited our searches to three databases: PubMed, CINHAL and Scopus. We conducted an extensive grey literature and snowball search by reviewing websites of numerous public health organisations and repositories, as well as emailing the authors’ respective networks. Supplementary File 1 provides a list of snowballing sources and completed database searches. Search terms were in both French and English. In addition, all included articles’ references were checked. To expedite the review process, two authors conducted the database search, three conducted grey literature and snowballing searches, and two conducted reference searching.

### Article Screening and extraction

All returned results were input into Covidence, a systematic review information management system, where duplicates were removed. The remaining articles were screened at title and abstract stage, and full-text stage independently by two reviewers, with a third resolving any discrepancies. Two team members independently screened all returned snowballing resources at full-text stage, with a third reviewer resolving any discrepancies. All authors participated in the screening.

Predefined and piloted data extraction tables were developed. Two authors initially extracted data from the included articles, with other authors reviewing all extractions for reliability and consistency. Content on community engagement actors/approaches and intervention focus was extracted directly as the articles reported if applicable, however this often did not occur leaving the review team to extrapolate and categorize. Given that research question seeks to identify what has been used, no quality ratings were applied to the included articles.

### Public and Patient Involvement (PPI)

There were no funds or time allocated for PPI so we were unable to involve patients. We encourage throughout the findings for programme and policy makers to involve communities within the design and implementation of their respective programmes.

## Results

Database and snowballing searches occurred between April 27^th^ and May 2^nd^, 2020. A total of 1112 articles were returned and after duplicate removal 956 abstracts were reviewed. In total 32 articles were identified for inclusion; five of which were identified through snowballing (four from initial grey literature/snowball search, and one from reviewing included articles’ references), and the remainder through database searches. Figure 1 presents the screening process and results.

**Figure 1:**
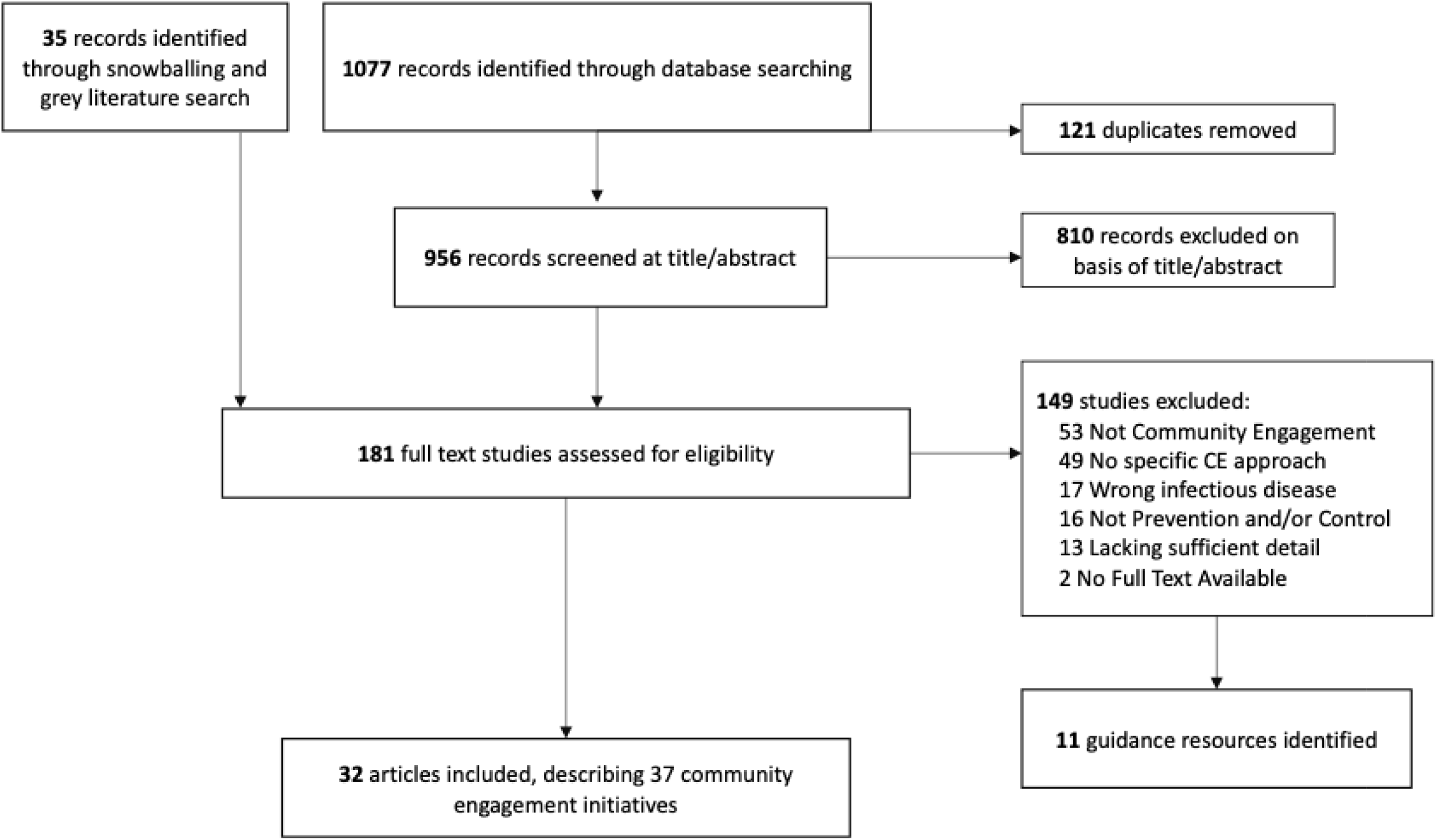
PRISMA Diagram.

In addition to the 32 documents included and reported within, 11 documents that did not address or describe a specific community engagement initiative but did provide overarching guidance to community engagement or aspects of community engagement, were identified. These documents were retained to provide additional implementation considerations for those undertaking community engagement. Box 1 provides a summary of these materials, and Supplementary File 3 includes additional details.

### Article Characteristics

Of the 32 included articles, all but three were published on or after 2015, with one article published in 2009 [31], one in 2010 [32] and one in 2012 [33]. The remaining were published in 2015 (n=2), 2016 (n=6), 2017 (n=9), 2018 (n=3), 2019 (n=3) and 2020 (n=6). All articles are in English, except for one which is in French [34]. 32 articles were included, however two articles report three [35], and four [36], distinct community engagement initiatives. As such, the remainder of the review will focus on 37 initiatives.

### Context and Outbreak

Of these 37 initiatives, 28 were for Ebola, with 25 relating to the 2014-2015 West Africa outbreak from Sierra Leone (n=11), Liberia (n=9), Guinea (n=2), Nigeria (n=1), Ghana (n=1) and one mixed-country study. The remaining three Ebola examples [37-39] were related to the 2018-2020 outbreak in the Democratic Republic of Congo, two of which focused on efforts in Uganda. Five community engagement initiatives were used for Zika within the United States and Puerto Rico (n=3), and one each in Singapore and Uruguay [40-43]. Four articles were specific to in H1N1, with three from Australia and one from Canada [31-33, 44].

Broad contextual concerns preceding the outbreak refer to poverty, unemployment or economic crisis [34, 45], health system failure, lack of development infrastructure [45, 46], colonial/post-colonial factors and ethnic and political conflicts [34, 35].

### Community Engagement Approaches and Interventions

The review identified 6 broad types of community engagement actors or approaches, which addressed infection prevention and control through six main channels. As highlighted in Table 2, the main approaches include: community leaders (traditional, religious and/or governing), community and faith-based organisations, community groups or networks or committees, health management committees, individuals (no further clarification provided) and key stakeholders which included students, survivors, women representatives, elderly and the youth. These community engagement interventions addressed infection prevention and control through six main channels: designing and planning interventions (including messaging), community entry and trust-building, social and behaviour change communication (SBCC), risk communication, surveillance and contract tracing, and broader logistics and administration activities such as procuring and setting-up hand washing stations, constructing facility or record keeping.

**Table 2:** Community engagement actors and their involvement in epidemic prevention and control activities

| Community Engagement Actors | Design and Planning | Community Entry/ Trust-Building | Social and Behaviour Change Communication | Risk Communication | Surveillance, Tracing | Logistics, Provision, Administration |
| --- | --- | --- | --- | --- | --- | --- |
| Leaders (traditional, religious and governing) | Charania and Tsuji, 2012; Juarbe-Rey et al. 2018; Miller et al. 2015; Kinsman et al. 2017 | Mbaye et al. 2017; Le Marcis et al. 2019; HC3, 2017a; Munodawafa et al. 2018; Skrip et al. 2020, | Gillespie et al. 2016; Barker et al. 2020; Mbaye et al. 2017; HC3, 2017a; HC3, 2017b; HC3, 2017c; HC3, 2017d; Aceng et al. 2020; Ho et al. 2017; Skrip et al. 2020; Gray et al. 2018; Jiang et al. 2016; Li et al. 2016 | Gillespie et al. 2016; Barker et al. 2020; Mbaye et al. 2017; Le Marcis et al. 2019a; ; Le Marcis et al. 2019c; HC3, 2017a; Aceng et al. 2020; ; Ho et al. 2017; Juarbe-Rey et al. 2018; Sepers et al. 2019; Skrip et al. 2020; Jiang et al. 2016; Li et al. 2016 | Barker et al. 2020; Mbaye et al. 2017; Le Marcis et al. 2019a; HC3, 2017a; HC3, 2017b; Aceng et al. 2020; Nakiire et al. 2020; Sepers et al. 2019; Gray et al. 2018; Li et al. 2017 | Barket et al. 2020; Gray et al. 2018; Le Marcis et al. 2019c |
|  | H1N1 (n=2), Zika (n=1), Ebola (n= 1) | Ebola (n=5) | Ebola (n= 12), Zika (n=1) | Ebola (n= 12), Zika (n=2) | Ebola (n= 10) | Ebola (n= 3) |
| Community-based organisations and faith organisations |  | Mbaye et al. 2017 | Mbaye et al. 2017; Santibañez et al. 2017 | Mbaye et al. 2017; Kirk-Sell et al. 2020; Adongo et al. 2016 | Mbaye et al. 2017 | Santibañez et al. 2017 |
|  |  | Ebola (n= 1) | Ebola (n= 1), Zika (n=1) | Ebola (n= 2), Zika (n=1) | Ebola (n= 1) | Zika (n= 1) |
| Community groups |  | Skrip et al. 2020 | HC3, 2017c Basso et al. 2017; Ho et al. 2017; Skrip et al. 2020; Gray et al. 2018; Abramowitz et al. 2017 | Le Marcis et al. 2019a; Ho et al. 2017; Skrip et al. 2020 | Le Marcis et al. 2019; Gray et al. 2018 | Gray et al. 2018 |
|  |  | Ebola (n= 1) | Ebola (n= 4), Zika (n=2) | Ebola (n= 2), Zika (n=1) | Ebola (n= 2) | Ebola (n= 1) |
| Health Management Committees/Community Health Committees |  |  | McMahon et al. 2017; Meredith, 2015 | McMahon et al. 2017; Meredith, 2015 | McMahon et al. 2017; Meredith, 2015 | McMahon et al. 2017; Meredith, 2015 |
|  |  |  | Ebola (n= 2) | Ebola (n= 2) | Ebola (n= 2) | Ebola (n= 2) |
| Individuals (Volunteers) | HC3, 2017c | Dada et al. 2019 | Barker et al. 2020; Aceng et al. 2020; Skrip et al. 2020; Jiang et al. 2016; Maduka et al. 2017 | Barker et al. 2020; Aceng et al. 2020; Skrip et al. 2020; Jiang et al. 2016; | Barker et al. 2020; Aceng et al. 2020; Nakiire et al. 2020; Ratnayake et al. 2016; Stone et al. 2016 | Barker et al. 2020 |

|  | Ebola (n= 1) | Ebola (n= 1) | Ebola (n= 5) | Ebola (n= 4) | Ebola (n= 5) | Ebola (n= 1) |
| --- | --- | --- | --- | --- | --- | --- |
| Key stakeholders | Massey et al. 2009; Rudge and Massey, 2010; Charania and Tsuji, 2012; Le Marcis et al. 2019b; Juarbe-Rey et al. 2018; Miller et al. 2015; Kinsman et al. 2017; | Massey et al. 2009 | Masumbuko et al. 2020; Ho et al. 2017; Gray et al. 2018 | Masumbuko et al. 2020; Ho et al. 2017; Juarbe-Rey et al. 2018; Li et al. 2016 | Li et al. 2016 |  |
|  | H1N1 (n=4), Zika (n=1), Ebola (n= 2) | H1N1 (n=1) | Ebola (n= 3), Zika (n=1) | Ebola (n= 2), Zika (n=2) | Ebola (n= 1) |  |
| **Totals** | 12 | 9 | 32 | 29 | 20 | 8 |

\*HC3 and Le Marcis have four and three examples of community engagement, respectfully. For the purpose of this table to demonstrate frequency of approaches, each example is cited as either a,b,c, or d. However, as these come from the same included article, references do not appear this way within the reference list

From Table 2, it can be seen that community engagement was mostly used for social and behaviour change communication and risk communication, followed by surveillance and contract tracing. Many of the reported community engagement activities involved multiple actors, and took multi-faceted approaches for prevention and control, as can be observed from Table 2. For example: Skrip et al. detail the Community-Led-Ebola-Action (CLEA) approach which involved community champions and mobilisers to support a structured participatory dialogue to identify and address community needs targeting areas of trust-building, risk communication and BCC [47]; McMahon et al. detail Health Management Committees, made up of leaders and key stakeholders, and their efforts in BCC and risk communication, and also supporting health facilities by conducting screening and administrative duties in relation to Ebola [48]; Ho and colleagues highlight how resident committees, grassroot leaders and volunteers conducted risk communication and source reduction for Zika [41]; and Mbaye and colleagues highlight how community groups, faith organisations and key stakeholders (youth, women and elderly) conducted trust-building, surveillance and SBCC [34].

The majority of the community engagement activities were not reported as a component of a larger programme, with the exception of surveillance systems which included community engagement for monitoring at the community level linked to a structured contract tracing system. Supplementary File 2 includes the extraction data for each article.

### Target groups and equity considerations

The majority of community engagement activities had community-wide focus, with no specific equity considerations reported. One article from Kirk-Sell et al. discusses community and faith-based organisations targeting marginalised populations, including non-English speakers and undocumented persons, in the US for risk communication in relation to Zika. On the contrary, all articles in relation to H1N1 had an equity focus; remote and isolated First Nations communities in Canada [33] and Aboriginal or Torres Strait Islanders communities in Australia [31, 32, 44]. Important to note however is that community engagement for these communities was limited to design and planning, with no reported inclusion of in implementation of activities.

Specific make-up of community engagement approaches was often not detailed or did not include diversity and representation though several reported community engagement structures including representation from Ebola survivors [50], women within reproductive age and students [42], women representatives [35], and youth [34, 47, 50].

### Health system linkages and support

Of those that provided detail on linkages, very few were explicitly linked to other health system components (with the exception of tracing). Community health committees [56] and Health Management Committees that were supporting health facility activities [48] were linked to Community Care Centres, and Ebola survivors, leaders and youth groups were used for behaviour change and surveillance, and linked with existing Community Health Workers [50].

### Implementation Considerations

Key implementation considerations for community engagement for COVID-19 prevention and control that were extracted from the included studies are presented in Figure 2. More broad implementation considerations synthesised from guidance documents (Supplementary File 3), are presented in Box 1. These considerations are also discussed in a Policy Brief on this research targeted towards implementers [61].

**Figure 2:**
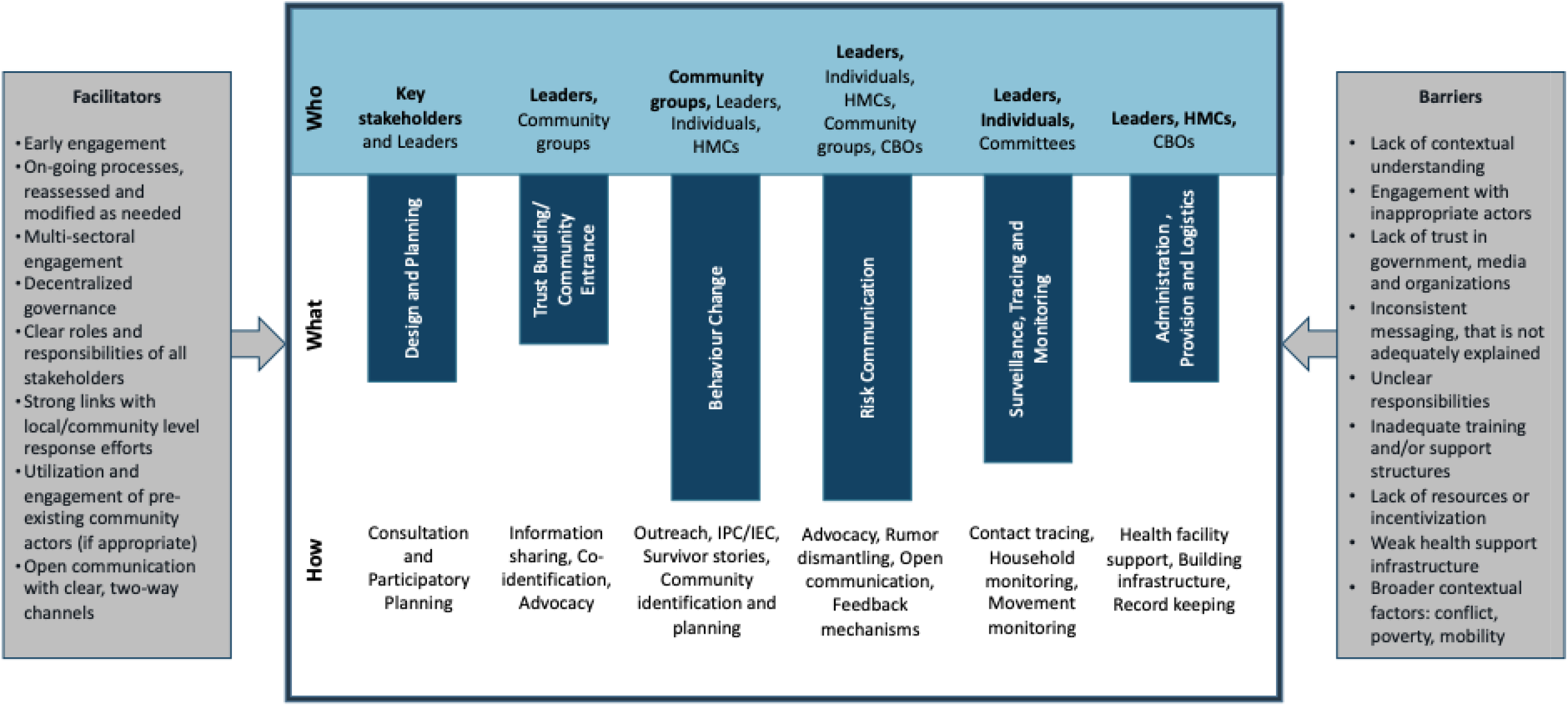
Components and Implementation Considerations of Community Engagement for Infectious Disease Prevention and Control. * The main CE actors (who) most common for that specific process are in bold. The length of the bars varies based on the most common way (what) of community engagement as per the reviewed literature. ‘How’ represents key activities that were undertaken within each broader intervention classification. **HMCs = Health Management Committees and include Community Health Committees; CFBOs = Community and Faith Based Organisations; IPC/IEC = Interpersonal Communication/Information, Education and Communication

**Box 1: Summary of implementation recommendations from guidance documents**

- Start early – community engagement should not be an afterthought, but fully incorporated into prevention and control activities
- Conduct contextual assessment which aims to identify cultures, traditions and customs (i.e. through anthropological research), and social norms, collective beliefs and barriers and facilitators (through social science methods)
- Engage local leadership (traditional and religious) and other key stakeholders early
- Collaborate with communities on issue identification and co-design of response
- Identify existing community engagement structures and resources and engage them (if appropriate)
- Utilize multi-sectoral action, and multi-level engagement
- Maintain two-way communication that is: clear, frequent, transparent, honest, positive focused (not fear inducing), respectful, tailored and piloted
- Incorporate feedback systems and regularly monitor and review initiatives when needed
- Support trust-building through collaborative engagement, maintaining expectations and promises, and providing inclusive, clear and honest information
- There is no ‘one-size-fits-all’ model for community engagement – different contexts require different approaches

## Discussion

Findings from this review highlight a need for more documentation of community engagement activities especially from more diverse geographic settings and across different populations. While some activities are underway, for instance GOAL Global, based on experience gained from their Ebola response, is implementing Community-Led Action (CLA) for COVID-19 in numerous countries [62], or Community Action Networks in Cape Town working together to identify and address the needs of community members [63], implementers, policy makers and researchers and encouraged to share learnings from past community engagement initiatives and document ongoing activities for COVID-19.

Interpretation of these findings should be done based on existing context, as the majority of articles were from Ebola response. Ebola had many unique considerations, including lack of trust, fear, rumours and cultural practices around burials [10]. Engagement of local leaders, those with high levels of respect, were critical to support dismantling some of these notions and working towards prevention and control activities. However, the COVID-19 response may parallel Ebola in many ways given the social spreading and potential stigma around contracting COVID-19.

Engagement lies on a spectrum, from more passive to active involvement. It can consist of providing *information* and conducting *consultation*, having *involvement* via regular interactions throughout the project cycle, to *collaboration* which entails working in partnership with shared decision-making [64, 65] that involves communities carrying out critical health systems functions and innovating with localised solutions [18]. Within this review, most included articles could be classified as having involvement, where communities were thoroughly brought in, but often did not share decision-making powers. Notably however, almost all examples of community engagement from high-income contexts consisted of consultation, demonstrating passive involvement with target ethnic and minority population. In addition, very few examples were identified that had an equity focus or strong equity considerations within target groups and engagement actors. While leadership buy-in is imperative for many community activities, so too is ensuring a balance between power and representation of diverse voices.

Community engagement may be specifically appropriate and needed for complex contexts, such as for migrants in humanitarian settings [66] or in urban informal settlements [67]. It is also needed to address more complex situations, such as settings dealing with both COVID-19 and risk of hunger [68] or supporting already overburdened health systems.

Countries with pre-existing community engagement structures can thoroughly and meaningfully embed such actions into national response plans. Recent modelling in Africa, where the large majority of articles including this review are based, has noted that if not controlled, COVID-19 could result in up to 190,000 deaths and 44 million infections in one year alone [69]. Many South Asian countries, which have recently seen exponential increases in COVID-19 cases, have a long history of community health and engagement activities and were some of the first to document the mobilization of community health workers like India’s ASHAs, for COVID-19. Countries without a strong history of community engagement need to identify where this may be most beneficial, for instance to support ethnic minorities in the global north who in many countries because of inequitable systems are being infected and killed at a disproportionate rate [70].

Community engagement supports shaping social dynamics based on power and control that perpetuate the marginalisation of certain groups. It needs to start early and continue after the critical stages of the health crisis to contribute to empowerment and building resilient communities. Addressing COVID-19 will require multi-sectoral responses and a variety of approaches from biomedical and social sciences. Community engagement should be a fundamental component within all these responses. Whether it be related to prevention and control, vaccine testing and ethics [57] or resilience and recovery [18], community engagement can support successful efforts. It can also have fundamental roles in rebuilding a stronger health system after the more acute phase of COVID-19 and supporting an equity-focused public health response. Though for all of these to work, community engagement needs to be meaningful, follow best practice recommendations and guidelines, and be specific to the context.

### Limitations

As this was a rapid review, our database searching and snowballing was limited in scope and time which may have resulted in missing articles. In addition, while our search terms attempted to include for all relevant topics related to community engagement, and we did include search terms for specific community-based interventions (i.e. SBCC and risk communication), this was not exhaustive which may have resulted in missing articles. Several articles were limited in detail, and extracting and labelling content was at the review team’s discretion, which may have resulted in incorrect coding on the type of actors and interventions. This may have been particularly relevant in situations where the engagement approaches and interventions conducted were of similar nature, for instance the distinction between CFBOs and community groups, and SBCC and risk communication. Nevertheless, this review shares important lessons regarding community engagement approaches from past epidemics that should guide COVID-19 response.

## Conclusion

COVID-19’s global presence and social transmission pathways require social and community responses. This may be particularly important to reach marginalised populations and support equity-informed responses. Previous experience from outbreaks shows that community engagement can take many forms and include different actors and approaches who support various prevention and control activities including design and planning, community entry and trust building, social and behaviour change communication, risk communication, surveillance and tracing, and logistics and administration. Countries worldwide are encouraged to assess existing community engagement structures and utilise community engagement approaches to support contextually specific, acceptable and appropriate COVID-19 prevention and control measures.

## Data Availability

All relevant data is included as supplementary files available upon request.

## Author Contribution

BG, SB and CAL proposed and designed the study. All authors contributed to searching, screening, extraction and synthesis of articles. ENM, RN and BG prepared the first draft of the manuscript, with all authors reviewing and providing inputs. All authors prepared and approved the final version of the manuscript. This research is a co-production of the Community Health – Community of Practice (CH-CoP) of which all authors are members.

## Acknowledgements

We would like to thank CH-CoP members and other networks who shared reports, guidelines and tools. We would like to thank Prof. Bruno Meessen and Hannah Sarah F. Dini for providing inputs into the protocol development and Jiawen Elyssa Liu for supporting dissemination.

## Abbreviations

CFBO: community and faith-based organisation
CLA: Community-led approach
CLEA: Community-Led-Ebola-Action
HMC: Health Management Committee
IEC: Information, Education and Communication
IPC: Interpersonal Communication
SBCC: Social and behavioural change communication

